# Reduced neutralizing activity of post-SARS-CoV-2 vaccination serum against variants B.1.617.2, B.1.351, B.1.1.7+E484K and a sub-variant of C.37

**DOI:** 10.1101/2021.07.21.21260961

**Authors:** Juan Manuel Carreño, Hala Alshammary, Gagandeep Singh, Ariel Raskin, Fatima Amanat, Angela Amoako, Ana Silvia Gonzalez-Reiche, Adriana van de Guchte, PARIS study group, Mahmoud Awawda, Radhika Banu, Katherine Beach, Carolina Bermúdez-González, Dominika Bielak, Liyong Cao, Rachel Chernet, Parnavi Desai, Shelcie Fabre, Emily. D. Ferreri, Daniel Floda, Charles Gleason, Hisaaki Kawabata, Zenab Khan, Giulio Kleiner, Denise Jurczyszak, Julia Matthews, Wanni Mendez, Lubbertus CF Mulder, Kayla Dr. Alberto E Paniz-Mondolfi, Russo, Ashley Salimbangon, Miti Saksena, A. Shin, Levy Sominsky, Johnston Tcheou, Komal Srivastava, Emilia Mia Sordillo, D. Noah Sather, Harm van Bakel, Florian Krammer, Viviana Simon

## Abstract

Highly efficacious vaccines against severe acute respiratory syndrome coronavirus 2 (SARS-CoV-2) have been developed.^1^ However, the emergence of viral variants that are more infectious than the earlier SARS-CoV-2 strains is concerning.^2^ Several of these viral variants have the potential to partially escape neutralizing antibody responses warranting continued immune-monitoring. Here, we tested a number of currently circulating viral variants of concern/interest, including B.1.526 (Iota), B.1.1.7+E484K (Alpha), B.1.351 (Beta), B.1.617.2 (Delta) and C.37 (Lambda) in neutralization assays using a panel of post-mRNA vaccination sera. The assays were performed with authentic SARS-CoV-2 clinical isolates in an assay that mimics physiological conditions. We found only small decreases in neutralization against B.1.526 and an intermediate phenotype for B.617.2. The reduction was stronger against a sub-variant of C.37, followed by B.1.351 and B.1.1.7+E484K. C.37 is currently circulating in parts of Latin America^3^ and was detected in Germany, the US and Israel. Of note, reduction in a binding assay that also included P.1, B.1.617.1 (Kappa) and A.23.1 was negligible. Taken together, these findings suggest that mRNA SARS-CoV-2 vaccines may remain effective against these viral variants of concern/interest and that spike binding antibody tests likely retain specificity in the face of evolving SARS-CoV-2 diversity.

## Introduction

Severe acute respiratory syndrome coronavirus 2 (SARS-CoV-2) emerged in late 2019 in China and has since then caused the coronavirus disease 2019 (COVID-19) pandemic. Vaccines were developed and distributed in record speed with emergency use authorizations as early as December 2020 (in the US). Coronaviruses are an exception among RNA viruses because their replication machinery possesses proofreading activity.^4^ It was, therefore, expected that they would evolve slower than other RNA viruses. The emergence of variant viruses, first in Europe in the summer of 2020^5-7^ - including in minks in Denmark - and then in late 2020 in the UK^8^, South Africa^9^ and Brazil^10^ was a surprising event. Since the detection of these first variants, several other variants of interest (VoIs) and variants of concern (VoCs) have spread locally as well as globally. These include B.1.526 (Iota) which emerged in New York City and comprises three different sub-lineages^11,12^, B.1.427/B.1.429 (Epsilon) which emerged in California,^13^ C.37 (Lambda) which emerged in Peru,^3^ B.1.617.2 (Delta) and B.1.617.1 (Kappa) which emerged in India,^14^ B.1.1.7+E484K (Alpha) which was detected in several countries, as well as A.23.1 which was first detected in Uganda.^15^ Several of these lineages are in decline, including B.1.526 and B.1.427/B.1.429 while others such as B.1.617.2 are increasing in prevalence. Many of these variants display extensive changes in the N-terminal domain (NTD) and/or the receptor binding domain (RBD) of spike, both of which harbor neutralizing epitopes. In addition, several of these variants also contain additional mutations in their spike gene that may enhance affinity to human angiotensin converting enzyme 2 (ACE2).^16^ These changes may impact negatively on binding and neutralization by vaccine-induced antibodies, which are discussed as mechanistic correlates of protection.^17,18^ It is, therefore, important to monitor neutralizing and binding activity of sera from vaccinees against these variants.

Here, we tested the neutralizing activity of a panel of 30 post mRNA SARS-CoV-2 vaccination sera against the three B.1.526 sublineages, B.1.1.7+E484K, B.1.617.2, B.1.351 and a C.37 sub-variant using a microneutralization assay. This assay was designed to allow for multicycle replication with sera/antibodies being present in the overlay at all times to better mimic physiological conditions.^19^ In addition, we performed binding assays against recombinant RBD and full-length spike proteins of several VoIs and VoCs.

## Results

First, we assessed the neutralizing activity of 30 post-vaccination serum samples from individuals vaccinated with mRNA-1273 (Moderna) or BNT162b2 (Pfizer/BioNTech). This panel of sera is used by the NIAID SARS-CoV-2 Assessment of Viral Evolution (SAVE) ‘*in vitro*’ group, which is part of the US Department of Health and Human Services (HHS) SARS-CoV-2 Interagency Group (SIG), to determine the immune escape of different VoCs and VoIs.^20^ The viruses tested in this study include the wild type WA1 isolate as well as clinical isolates derived from nasopharyngeal swabs obtained from patients diagnosed with COVID-19 at the Mount Sinai Health System in New York City, representing B.1.526 (no change in RBD), B.1.526 (E484K), B.1.526 (S477N), B.1.1.7+E484K, B.1.351, B.1.617.2 and a subvariant of C.37 (**Supplementary Table 1, Supplemental Figure 1**). The C.37 subvariant contained additional changes compared to the consensus sequence including a large 13 amino acid deletion instead of G75V and T76I NTD substitutions and an additional E471Q substitution in the RBD. The panel of post-vaccination sera (**Supplementary Table 2**) was tested against these seven viruses in a well-established neutralization assay.^19^ In this assay, 60 tissue culture infectious doses of virus per well are pre-incubated with the respective serum dilutions for one hour. The serum/virus mix is then used to infect Vero.E6 cells. After the infection step, the cell culture medium is replaced with fresh medium containing the respective concentration of serum. The assay allows, therefore, for multicycle replication of the test virus in the continuous presence of test serum, which recapitulates infection conditions *in vivo*.

The microneutralization assays showed only modest reductions in neutralizing activity against the three B.1.526 variants compared to the wild type WA1 isolate (**Figure 1**). The geometric mean titer (GMT) of neutralization of the three B.1.526 sublineages was reduced by 1.4-fold (ancestral B.1.526), 1.8-fold (E484K) and 2.3-fold (S477N), respectively. Reduction in neutralizing activity against the B.1.617.2 isolate was moderate with 3-fold change compared to WA1. A more pronounced drop in neutralizing activity was seen with the B.1.1.7+E484K isolate (3.8-fold). As expected and reported before, an even stronger decrease in neutralization (4.2-fold) was found against the B.1.351 isolate. However, the greatest reduction in GMT (4.6-fold) was observed against a C.37 variant isolate. Of note, as explained above, virus isolate PV29369 carries additional substitutions/deletions in spike (see **Supplemental Table 1**) compared to the majority of circulating C.37.

**Figure 1.**
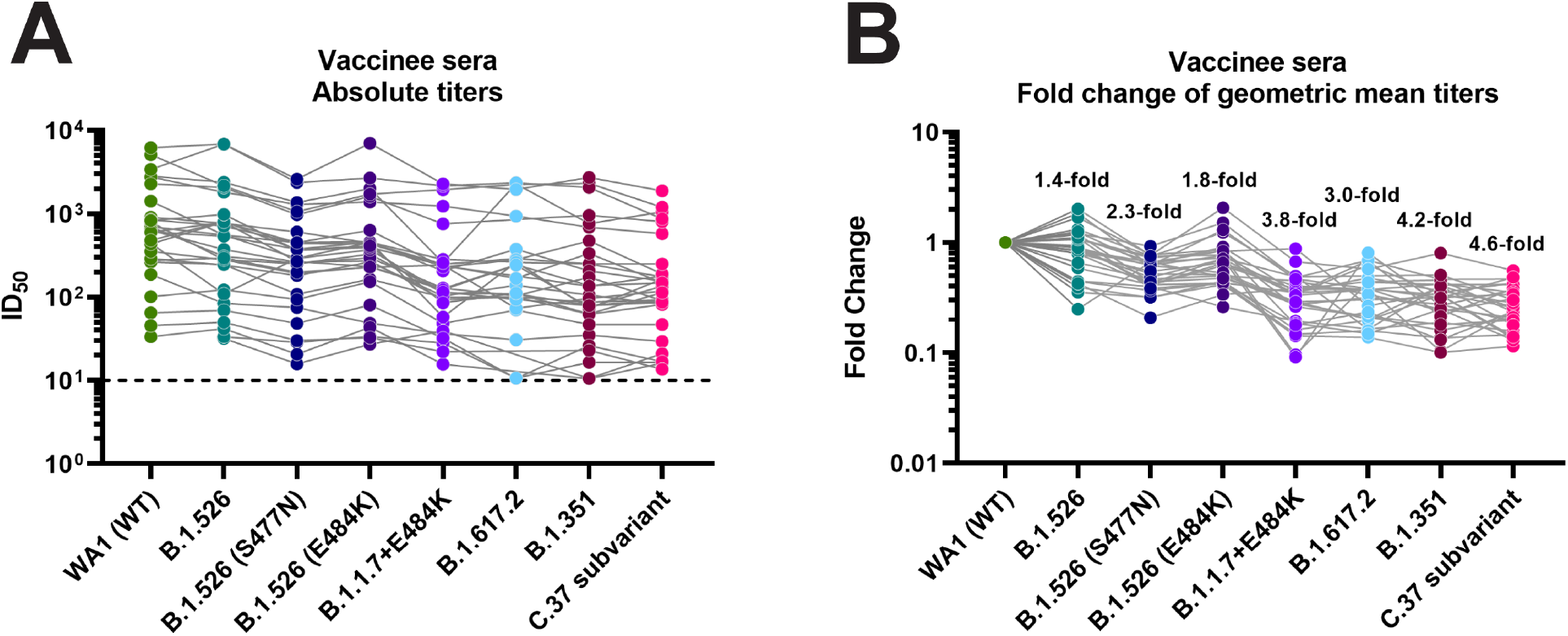
Neutralizing activity of post-mRNA vaccination sera against different VoIs and VoCs. **A** Absolute neutralization titers against the respective variant viruses. **B** Fold reduction of neutralization as compared to the wild type WA1 isolate. A list of specific mutations of the clinical isolates can be found in Supplementary Table 1.

We next determined antibody-binding activity against variant RBDs and spike proteins. The fold-reduction in binding against variant RBDs from B.1.1.7, P.1 and B.1.351 was negligible with a maximum reduction of 1.6-fold against the B.1.351 RBD (**Figure 2A and B**). Similarly, reduction in binding to B.1.1.7, B.1.351. P.1, B.1.617.1 and A.23.1 spike proteins was minimal, again with B.1.351 showing the strongest impact and a 1.4-fold reduction (**Figure 2C and D**).

**Figure 2.**
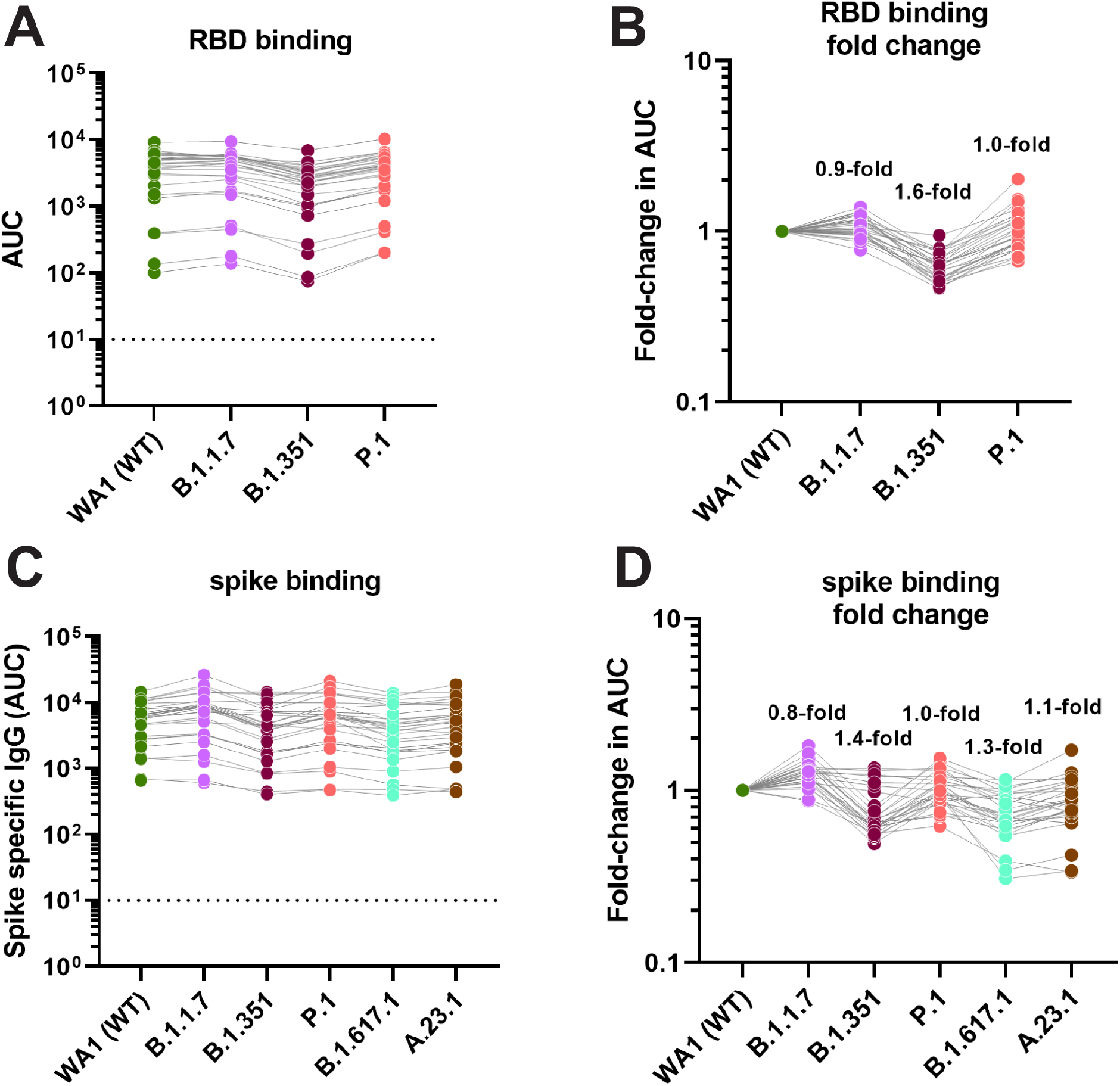
Binding activity of post-mRNA vaccination sera against RBD and spike proteins of different VoIs and VoCs. **A** and **C** show absolute area under the curve (AUC) binding values against various RBD and spike proteins, **B** and **D** show fold-reduction in binding as compare to Wuhan-1 derived wild type proteins.

## Discussion

Monitoring the effectiveness of vaccination in the context of evolving viral variants is of great public health concern for control of the COVID-19 pandemic. Indeed, neutralizing antibodies play a major role in protection from COVID-19, as shown both in animal models as well as in humans.^17,18,21-23^ It is, therefore, important to continuously monitor emerging variants and their impact on neutralizing activity of post-vaccination serum. Our data show a spectrum of neutralizing activities against a panel of VoIs and VoCs. The least negative impact on neutralization was seen for B.1.526 variant, which is reasonable since this isolate has very few changes in the spike protein (T95I, D253G and D614G) compared to the wild type WA1 isolate. The impact was higher for the B.1.526 isolates that carry, in addition, E484K or S477N substitutions in the RBD. Interestingly, S477N exerted a stronger impact than E484K, which was surprising given the strong reduction caused by introduction of E484K into the WA1 wild type backbone.^24^ However, S477N has also been described as escape mutation.^25^ The reduction in neutralization against B.1.617.2 was moderate, which aligns well with the preserved vaccine effectiveness against this variant.^26,27^ The combination of B.1.1.7 (which already harbors two deletions in the NTD) with E484K led to a substantial reduction in neutralizing activity. However, although B.1.1.7+E484K emerged several times independently, it does not seem to spread well – potentially due to the detrimental effect that the E484K mutation can have on binding affinity to human ACE2.^16^ As expected and already shown by several publications,^28-33^ neutralizing activity to B.1.351 was strongly reduced likely due to the three changes in RBD and extensive changes in the NTD. Interestingly, neutralizing activity was even more reduced for the local isolate of C.37 used in the current study which has more extensive changes in the NTD (Δ63-75, Δ246-252) and the RBD (L452Q, E471Q, F490S) compared to the predominant C.37 variant (NTD: G75V, T76I, Δ246-252; RBD: L452Q, F490S).^3^ While there was a strong reduction in neutralizing activity of post-vaccination sera against this variant, all 30 sera retained at least partial neutralization activity against this C37 variant virus which may indicate that – similar to B.1.351^34^ – mRNA vaccines will remain effective against these VoIs. Of note, the additional changes in the local C.37 isolate likely contribute to the immune evasion phenotype shown here and should be seen as worst case scenario for the C.37 variant.

Another interesting finding was that the reduction in binding activity to variant RBD and spike proteins was negligible. However, while neutralization activity is mostly dependent on only a few epitopes in the RBD and NTD, many more binding epitopes exist on the spike,^16^ which may explain our findings. Interestingly, a recent study found even better correlation between binding activity and vaccine effectiveness than for neutralizing activity and vaccine effectiveness.^17^ This may indicate that binding antibodies – even if they lack neutralizing activity – are involved in protection. It is well known that non-neutralizing antibodies raised upon infection or vaccination by ebolavirus or influenza viruses, can be protective via Fc-mediated effector functions.^35-38^ Future studies are needed to establish if this is also the case for SARS-CoV-2. However, the finding that the drop in binding activity is negligible even against spike proteins from variants with extensive changes like B.1.351 is reassuring in that context.

## Materials and Methods

### Cells and SARS-CoV-2 isolates

Vero.E6 cells were cultured in Dulbecco’s Modified Eagles Medium (DMEM, Corning) containing 10% fetal bovine serum (FBS, Corning) supplemented with 100 U/ml penicillin, 100 μg/ml streptomycin (Gibco). Nasopharyngeal swab specimens were collected as part of the routine SARS-CoV-2 surveillance conducted by the Mount Sinai Pathogen Surveillance program (IRB approved, HS#13-00981). Specimens were selected for viral culture on Vero-E6 cells based on the complete viral genome sequence information.^39^ Viruses were grown in Vero.E6 cells for 4-6 days; the supernatant was clarified by centrifugation at 4,000 g for 5 minutes and aliquots were frozen at -80°C for long term use. Expanded viral stocks were sequence-verified and titered on Vero.E6 cells prior to use in microneutralization assays.

### Phylogeny of SARS-CoV-2 lineages

The phylogenetic relationships of the SARS-CoV-2 isolates from the Pathogen Surveillance Program of the Mount Sinai Health System are depicted in a New York focused background from GISAID. The timed tree was built with nextstrain.org/sars-cov-2. PANGO lineages are depicted for the VoIs and VoCs used in the neutralization assays.

### Post vaccination serum samples

A panel of 30 post-vaccination sera was generated for use and distribution by BEI Resources for the NIAID SAVE “*in vitro*” group (**Supplementary Table 2**). Briefly, 30 participants enrolled in the PARIS (Protection Associated with Rapid Immunity to SARS-CoV-2) study (reviewed and approved by the Mount Sinai Hospital Institutional Review Board; IRB-20-03374) were invited to provide additional serum samples after receiving their second mRNA vaccine dose. All specimens were coded prior to processing, banking and sharing with BEI Resources. 15 participants had received two doses of the mRNA-1273 (Moderna) and 15 participants had received two doses of the BNT162b2 (Pfizer/BioNTech). The majority of participants were naïve prior to vaccination with 4-5 COVID-19 survivors being included each vaccine group.

### ELISA

Antibody titers in sera from vaccines were determined by a research grade enzyme-linked immunosorbent assay (ELISA) using recombinant versions of the receptor binding domain (RBD) and the spike (S) of wild type SARS-CoV-2 and variant viruses (RBD: WT/Wuhan-1, B.1.1.7, B.1.351, P.1; spike: WT/Wuhan-1, B.1.1.7, B.1.351, P.1, B.1.617.1, A.23.1). Briefly, 96-well microtiter plates (Thermo Fisher) were coated with 50 μl/well of recombinant protein (2 μg/ml) overnight at 4 °C. Plates were washed three times with phosphate-buffered saline (PBS; Gibco) supplemented with 0.1% Tween-20 (PBS-T; Fisher Scientific) using an automatic plate washer (BioTek). For blocking, PBS-T containing 3% milk powder (American Bio) was used. After 1-hour incubation at room temperature (RT), blocking solution was removed and initial dilutions of heat-inactivated sera (in PBS-T 1%-milk powder) were added to the plates, followed by 2-fold serial dilutions. After 2-hour incubation, plates were washed three times with PBS-T and 50 μl/well of the pre-diluted secondary antibody anti-human IgG (Fab-specific) horseradish peroxidase antibody (produced in goat; Sigma, A0293) diluted 1:3,000 in PBS-T containing 1% milk powder were added. After 1-hour incubation at RT, plates were washed three times with PBS-T and SigmaFast o-phenylenediamine dihydrochloride (Sigma) was added (100 μl/well) for 10min, followed by addition of 50 μl/well of 3 M hydrochloric acid (Thermo Fisher) to stop the reaction. Optical density was measured at a wavelength of 490 nm using a plate reader (BioTek). Area under the curve (AUC) values were calculated and plotted using GraphPad Prism 7.

### Microneutralization assay

Sera from vaccinees were used to assess the neutralization of wild type SARS-CoV-2 and selected VoIs and VoCs (Supplementary **Table 1**). All procedures were performed in the Biosafety Level 3 (BSL-3) facility at the Icahn School of Medicine at Mount Sinai following standard safety guidelines. Remdesivir was used as a control for wild type and variant viruses to monitor assay variation. The day before infection, Vero E6 cells were seeded in 96-well high binding cell culture plates (Thermo Fisher) at a density of 20,000 cells/well in complete Dulbecco’s Modified Eagle Medium (cDMEM). After heat inactivation of sera (56°C for 1 hour), serum samples were serially diluted (3-fold) in minimum essential media starting at 1:10. For dilutions of sera, infection media consisting of minimum essential media (MEM, Gibco) supplemented with 2 mM L-glutamine, 0.1% sodium bicarbonate (w/v, Gibco), 10 mM 4-(2-hydroxyethyl)-1-piperazineethanesulfonic acid (HEPES, Gibco), 100 U/ml penicillin, 100 μg/ml streptomycin (Gibco) and 0.2% bovine serum albumin (MP Biomedicals) was used. Vero E6 cells were seeded in 96-well high binding cell culture plates (Thermo Fisher) at a density of 20,000 cells/well in complete Dulbecco’s Modified Eagle Medium (cDMEM). On the next day, sera dilutions were incubated with 1,000 tissue culture infectious dose 50 (TCID50) of wt USA-WA1/2020 SARS-CoV-2 or the corresponding variant virus for 1 hour at RT, followed by transfer of 120μl of virus-sera mix to Vero-E6 plates. Infection was led to proceed for 1 hour at 37°C and inoculum was removed. 100 μl/well of the corresponding antibody dilutions plus 100μl/well of infection media supplemented with 2% fetal bovine serum (Gibco) were added to the cells. Plates were incubated for 48h at 37°C.

For staining of the nucleoprotein antigen, cells were fixed by addition of 150 μl/well of a 10% formaldehyde solution and incubated overnight at 4°C. Fixed cell monolayers were washed with PBS (Gibco) and permeabilized with 150 μl/well of PBS, 0.1% Triton X-100 for 15 min at RT. Permeabilization solution was removed, plates were washed with 200 μl/well of PBS twice and 200 μl/well of blocking solution consisting of PBS 3% milk (American Bio) were added for 1 h at RT. Blocking solution was removed and 100 μl/well of mAb 1C7 (anti-SARS nucleoprotein antibody produced in house) diluted in PBS 1% milk (American Bio) at a 1:4000 dilution were added for 1 h at RT. Primary antibody was removed and plates were washed with 200 μl/well of PBS twice, followed by addition of 100 μl/well of goat anti-mouse IgG-HRP (Rockland Immunochemicals) diluted in PBS 1% milk (American Bio) 1:3000. After 1 hour incubation, antibody solution was removed, plates were washed twice with PBS and 100 μl/well of o-phenylenediamine dihydrochloride (Sigmafast OPD; Sigma-Aldrich) were added for 10 min at RT. The reaction was stopped by addition of 50 μl/well of HCl 3M (Thermo Fisher Scientific). Optical density (OD) was measured (490 nm) with a microplate reader (Synergy H1; Biotek). Initial subtraction of background and calculation of the percentage of neutralization with respect to the “virus only” control was done in excel. A non linear regression curve fit analysis, using Prism 9 software (GraphPad), was performed to calculate the inhibitory dilution 50% (ID50), using top and bottom constraints of 100% and 0% respectively.

## Data Availability

All data referred to in the manuscript are available

## Acknowledgments

We would like to thank the study participants for their generosity and willingness to participate in longitudinal COVID19 research studies. None of this work would be possible without their contributions. We would like to thank Dr. Randy A. Albrecht for oversight of the conventional BSL3 biocontainment facility, which makes our work with live SARS-CoV-2 possible. We are also grateful for Mount Sinai’s leadership during the COVID-19 pandemic. We want to especially thank Drs. Peter Palese, Carlos Cordon-Cardo, Dennis Charney, David Reich and Kenneth Davis for their support. This work is part of the PARIS/SPARTA studies funded by the NIAID Collaborative Influenza Vaccine Innovation Centers (CIVIC) contract 75N93019C00051. This work was also partially funded by the Centers of Excellence for Influenza Research and Surveillance (CEIRS, contract # HHSN272201400008C), and by the generous support of the JPB Foundation and the Open Philanthropy Project (research grant 2020-215611 (5384); and by anonymous donors. Finally, this effort was also supported by the Serological Sciences Network (SeroNet) in part with Federal funds from the National Cancer Institute, National Institutes of Health, under Contract No. 75N91019D00024, Task Order No. 75N91020F00003. The content of this publication does not necessarily reflect the views or policies of the Department of Health and Human Services, nor does mention of trade names, commercial products or organizations imply endorsement by the U.S. Government.

## Figure Legends

**Supplementary Table 1.**
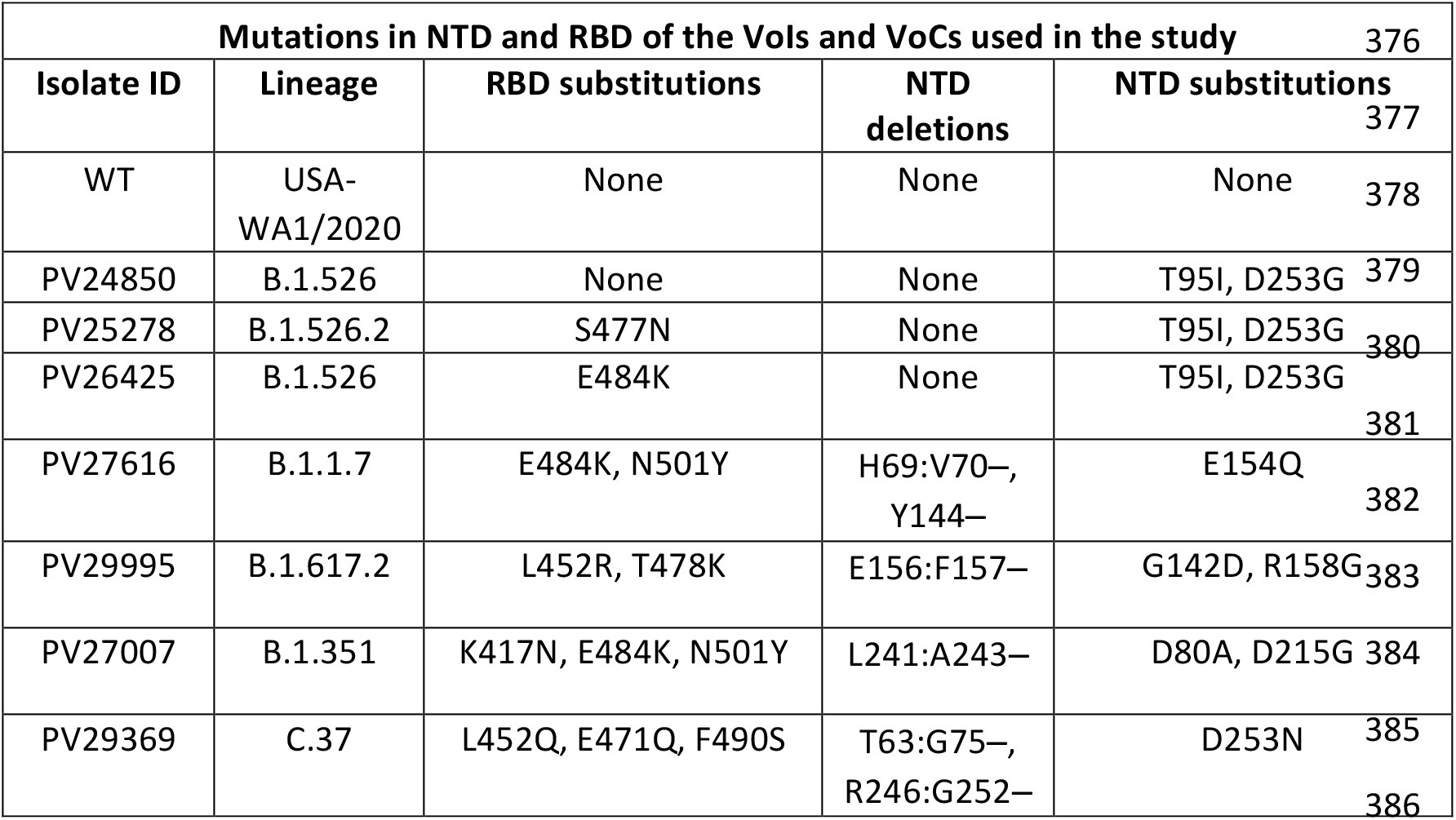
Information of viral isolates used in microneutralization assays.

**Supplementary Table 2.** Sera used in the neutralization assays.

| # | sample | Vaccine # | Serostatus* | Sex | Days after 1st dose | Days after 2<sup>nd</sup> dose |
| --- | --- | --- | --- | --- | --- | --- |
| 1 | 77579 | Moderna | neg | Female | 30 | 2 |
| 2 | 76350 | Moderna | pos | Male | 35 | 7 |
| 3 | 26973 | Moderna | neg | Female | 32 | 5 |
| 4 | 95482 | Moderna | neg | Female | 31 | 3 |
| 5 | 93764 | Moderna | neg | Female | 34 | 9 |
| 6 | 38258 | Moderna | neg | Male | 34 | 5 |
| 7 | 14275 | Moderna | pos | Female | 30 | 2 |
| 8 | 22686 | Moderna | pos | Female | 35 | 5 |
| 9 | 80902 | Moderna | neg | Female | 32 | 6 |
| 10 | 91353 | Moderna | pos | Female | 32 | 5 |
| 11 | 66902 | Moderna | neg | Female | 35 | 10 |
| 12 | 47380 | Moderna | neg | Female | 37 | 9 |
| 13 | 32885 | Moderna | neg | Male | 31 | 3 |
| 14 | 30756 | Moderna | pos | Female | 31 | 3 |
| 15 | 72329 | Moderna | neg | Male | 33 | 6 |
| 16 | 17146 | Pfizer | neg | Female | 47 | 22 |
| 17 | 35098 | Pfizer | neg | Male | 20 | 6 |
| 18 | 94525 | Pfizer | neg | Male | 28 | 7 |
| 19 | 70771 | Pfizer | neg | Male | 48 | 27 |
| 20 | 13967 | Pfizer | neg | Female | 29 | 8 |
| 21 | 29021 | Pfizer | neg | Female | 42 | 23 |
| 22 | 11348 | Pfizer | neg | Male | 43 | 22 |
| 23 | 67169 | Pfizer | neg | Female | 46 | 22 |
| 24 | 91699 | Pfizer | pos | Female | 49 | 29 |
| 25 | 99695 | Pfizer | pos | Male | 43 | 21 |
| 26 | 58441 | Pfizer | pos | Male | 43 | 25 |
| 27 | 79044 | Pfizer | pos | Female | 43 | 22 |
| 28 | 87845 | Pfizer | neg | Female | 39 | 20 |
| 29 | 48139 | Pfizer | neg | Male | 29 | 9 |
| 30 | 26461 | Pfizer | neg | Female | 43 | 21 |

\* Serostatus denotes SARS-CoV-2 antibodies prior to COVID19 vaccination

**Supplementary Figure 1:**
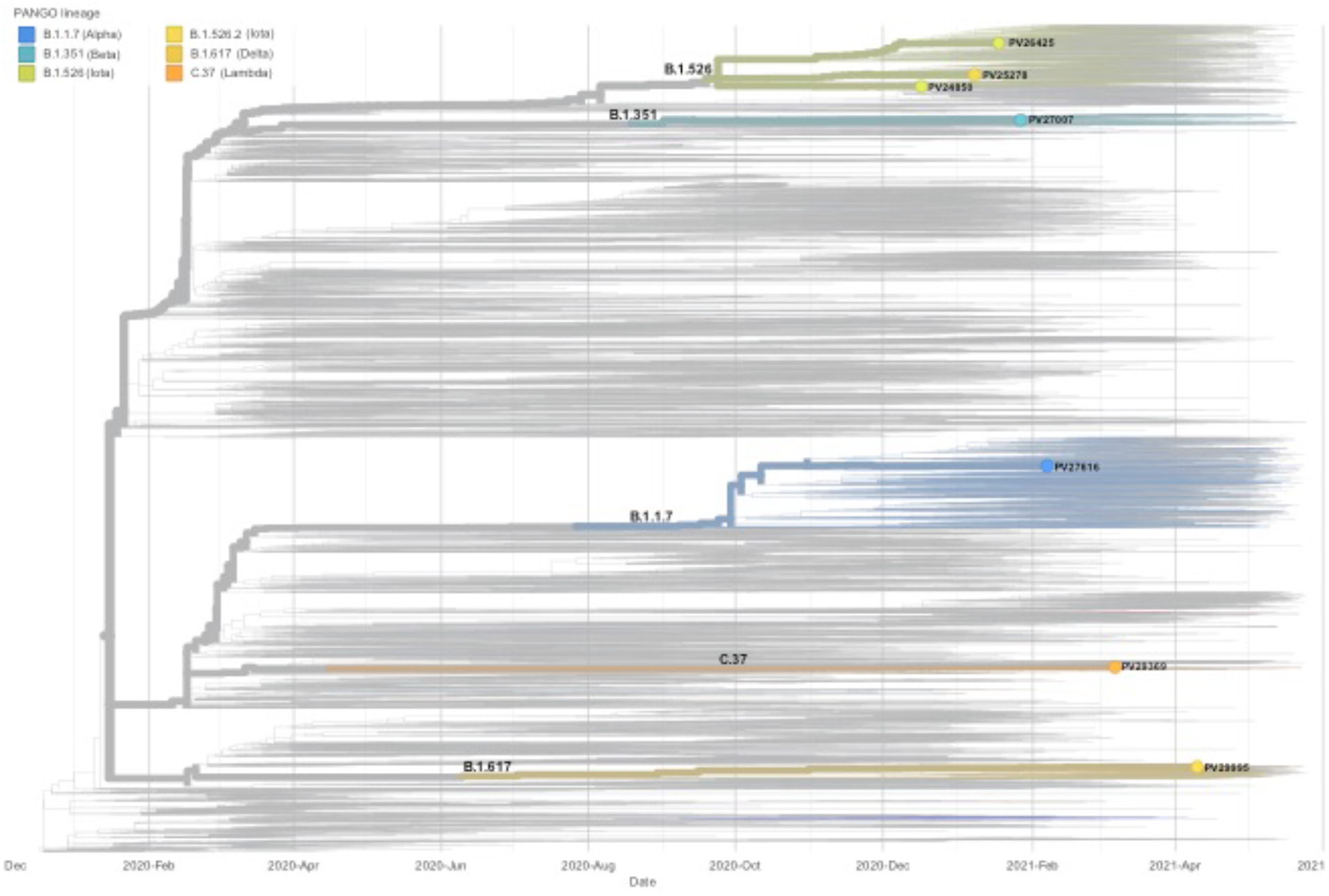
Phylogenetic tree including all patient isolates used in this study.

